# Lockdowns exert selection pressure on overdispersion of SARS-CoV-2 variants

**DOI:** 10.1101/2021.06.30.21259771

**Authors:** Bjarke Frost Nielsen, Andreas Eilersen, Lone Simonsen, Kim Sneppen

**Affiliations:** Niels Bohr Institute, University of Copenhagen, Blegdamsvej 17, 2100 Copenhagen, Denmark; Department of Science and Environment, Roskilde University, Universitetsvej 1, 4000 Roskilde, Denmark

**Keywords:** Overdispersion, Evolution, Superspreading, Non-pharmaceutical interventions

## Abstract

The SARS-CoV-2 ancestral strain has caused pronounced super-spreading events, reflecting a disease characterized by overdispersion, where about 10% of infected people causes 80% of infections. New variants of the disease have different person-to-person variations in viral load, suggesting for example that the Alpha (B.1.1.7) variant is more infectious but relatively less prone to superspreading. Meanwhile, mitigation of the pandemic has focused on limiting social contacts (lockdowns, regulations on gatherings) and decreasing transmission risk through mask wearing and social distancing. Using a mathematical model, we show that the competitive advantage of disease variants may heavily depend on the restrictions imposed. In particular, we find that lockdowns exert an evolutionary pressure which favours variants with lower levels of overdispersion. We find that overdispersion is an evolutionarily unstable trait, with a tendency for more homogeneously spreading variants to eventually dominate.

**Significance:** One of the most important and complex properties of viral pathogens is their ability to mutate. The SARS-CoV-2 pandemic has been characterized by overdispersion – a propensity for superspreading, which means that around 10% of those who become infected cause 80% of infections. However, evidence is mounting that this is not a stable property of the virus and that the Alpha variant spreads more homogeneously. We use a mathematical model to show that lockdowns exert a selection pressure, driving the pathogen towards more homogeneous transmission. In general, we highlight the importance of understanding how non-pharmaceutical interventions exert evolutionary pressure on pathogens. Our results imply that overdispersion should be taken into account when assessing the transmissibility of emerging variants.

---

**O**ne of the major features of the coronavirus pandemic has been overdispersion in transmission, manifesting itself as superspreading. There is evidence that around 10% of infected individuals are responsible for 80% of new cases (1– 4). This means that some individuals have a high personal reproductive number, while the majority hardly infect at all. A recent study has shown this is reflected in the distribution of viral loads which is extremely wide, with just 2% of of SARS-CoV-2 positive individuals carrying 90% of the virus particles circulating in communities (5). Overdispersion is in fact a key characteristic of certain diseases (6–8). However, this is by no means a universal signature of infectious respiratory diseases. Pandemic influenza, for example, is characterized by a much more homogeneous transmission pattern (9–11).

As an emerging virus evolves, its transmission patterns may change and it may become more or less prone to superspreading. The Alpha (B.1.1.7) variant of SARS-CoV-2 has been reported to be ∼ 50% more transmissible than the ancestral SARS-CoV-2 virus under varying degrees of lockdown (12–14). Meanwhile, others have shown that the Alpha variant possesses a higher *average* viral load and a reduced variability between infected persons, compared to the ancestral strain (15, 16). It remains to be seen how this reduced variability affects the transmission patterns of the virus.

The altered viral load distributions seen in persons infected with the Alpha variant have also been investigated at the level of individual mutations. The spike protein of the Alpha variant prominently features the N501Y substitution (asparagine replaced by tyrosine at the 501 position) as well as the ΔH69/V70 deletion (histidine and valine deleted at the 69 and 70 positions). Investigators found that the viral load is, on average, three times as great for the Alpha variant compared with the ancestral strain (16). Furthermore, viral load distributions in samples taken from persons infected with a variant with the ΔH69/V70 show a lower variance, whether or not they also have tyrosine at the 501 position. However, the difference in variance was most pronounced for those samples which had the deletion as well as the 501Y mutation. Similarly, an analysis of samples with the N501Y mutation show that they have a higher median viral load as well as a substantially diminished variance compared to those without it. Using data from Ref. (15), we calculate that the viral loads in samples of the Alpha variant are associated with a lower coefficient of variation of approximately 2, compared to 4 for the ancestral strain. Importantly, the exact relation between viral load and infectiousness is not well understood; however, a higher viral load is logically expected to increase the risk of disease transmission. By this logic, the decreased variability in the viral load for the Alpha variant may translate into a reduced overdispersion in transmission.

In this paper, we use a mathematical model to study the competition between idealized variants which differ in their level of overdispersion (*k*) and their mean infectiousness. Our focus is on exploring whether overdispersion confers any evolutionary (dis)advantages, and whether non-pharmaceutical interventions which restrict social network size and transmissibility change the fitness landscape for variants with varying degrees of overdispersion. While it is evident that a higher mean infectiousness confers an evolutionary advantage to an emerging pathogen, it is not *a priori* obvious if a competitive advantage can be gained by specifically altering the *variability* in infectiousness (while keeping transmissibility unchanged). Our recent studies have shown that the presence of overdispersion makes a pandemic far more controllable than influenza pandemics when mitigating by limiting non-repetitive contacts (17) and personal contact network size (18). We therefore speculate that restrictions which alter social contact structure may, conversely, provide a fitness advantage to variants with more homogeneous transmission, and may thus play a role in viral evolution.

Across several diseases, individual variations in infectiousness have been approximated by a Gamma distribution (6) characterized by a certain mean value and a dispersion parameter known as *k*, which is rel<u>a</u>ted to the coefficient of variation (*CV*) through 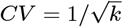. In the simplest of cases (a well-mixed population), infection attempts are modeled as a constant-rate (Poisson) process, which leads to a personal reproductive number which follows a negative binomial distribution. The dispersion parameter *k* characterizes the degree of transmission heterogeneity; a *lower k* corresponds to greater heterogeneity. For small values of *k*, it approximately corresponds to the fraction of infected individuals responsible for 80% of new infections The value for the SARS-CoV-2 ancestral virus is around 10%, corresponding to a *k*-value of approximately 0.1. Other coronaviruses are also prone to superspreading, with the *k*-values of SARS-CoV-1 and MERS estimated at 0.16 (6) and 0.26 (19), respectively. To explore questions of how such overdispersion affects fitness and pathogen evolution, we use an agent-based model of COVID-19 spreading in a social network, as originally developed in Ref. (18).

Overdispersion in personal reproductive number – i.e. superspreading – is a phenomenon that requires *means* (biological infectiousness) as well as *opportunity* (social context). Superspreading can have diverse origins, ranging from purely behavioural to biological (8, 20). However, a recent meta-review (21) compared the transmission heterogeneity of influenza A (H1N1), SARS-CoV-1 and SARS-CoV-2 and found that higher variability in respiratory viral load was closely associated with increased transmission heterogeneity. This suggests that biological aspects of individual diseases are decisive in determining the level of overdispersion, and thus the risk of superspreading.

## Initial survival of variants

The words *fitness* and *competitive advantage* may take on several meanings in an evolutionary context. For our purposes, it is especially important to distinguish between the ability of a pathogen to *avoid stochastic extinction* and to *reproduce effectively* in a population.

To quantify the ability to avoid stochastic extinction we use a branching process to simulate an outbreak of a variant with a given level of overdispersion in a naive population. We then record whether it survives beyond the first 10 generations of infections, as a measure of the ability of that variant to take hold. Repeating these simulations multiple times allows us to compute the survival chance of each variant as a function of its infectiousness and overdispersion, in the absence and presence of mitigation (Fig. 1). Since we are dealing with a few related quantities, some definitions must be made. By the *basic reproductive number* (*R*_0_) we mean the average number of new infections which each infected person gives rise to *when all contacts are susceptible*. This is in contrast to the effective reproductive number (known variously as *R, R*_*t*_ and *R*_*e*_), which is affected by population immunity. Note that *R*_0_ as well as *R*_*e*_ are context dependent, since behaviour (and mitigation strategies) will affect e.g. the number of contacts that a person has and thus the reproductive number. Another parameter entirely is the *(biological) mean infectiousness*, by which we mean the rate at which transmission occurs *when* an infected person is in contact with a susceptible person. This is a property of the disease and not of the social environment. In Fig. 1, the independent variables are thus the mean infectiousness and the dispersion parameter, both of which are assumed to be properties of the disease. The details of the calculation can be found in the Materials and Methods section.

**Fig. 1.**
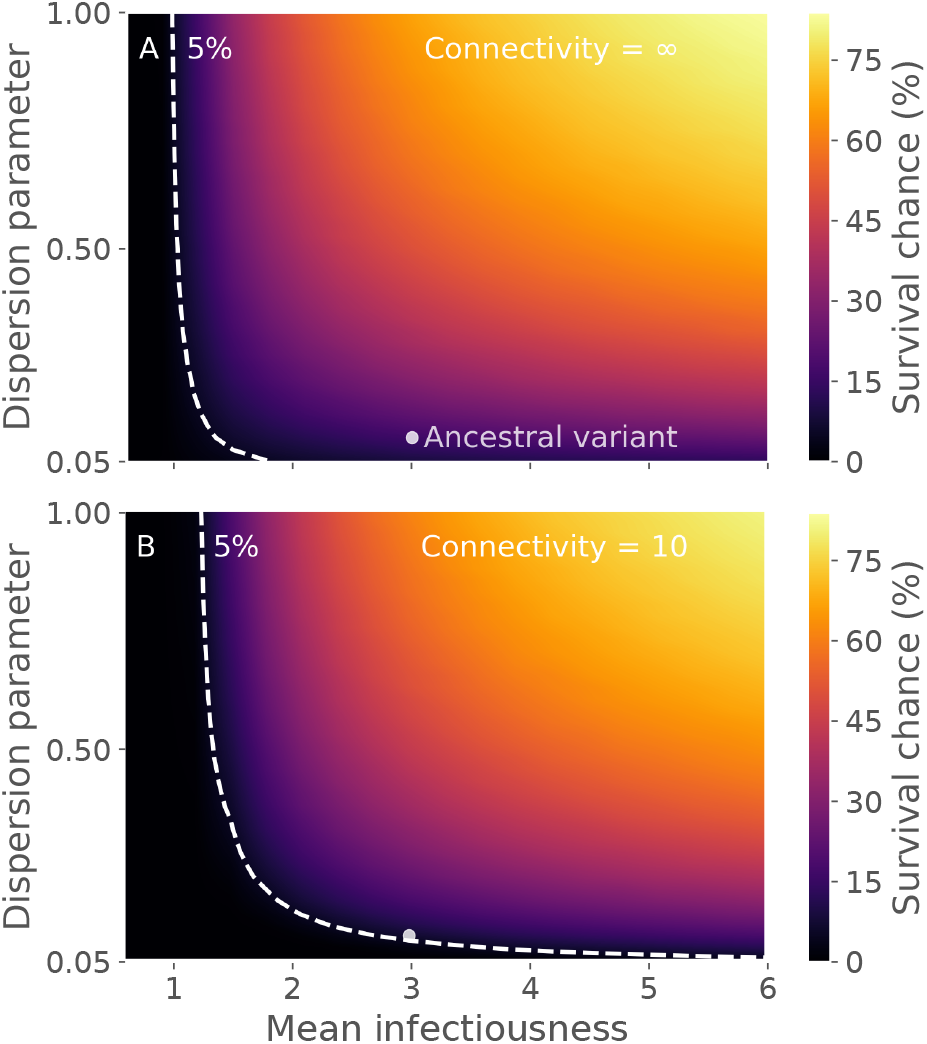
Initial survival chance depends strongly on overdispersion and moderately on lockdown status. **A)** The epidemic spreads in an unrestricted setting (homogeneous mixing contact structure) **B)** The epidemic spreads in a situation with limited social connectivity (modeled as an Erdos-Renyi network of average connectivity 10). The survival chance is computed by simulating several outbreaks, each starting from a single infected individual in a susceptible population. This initial individual is infected with a variant of a given overdispersion. For each outbreak, the variant is recorded as having *survived* if it does not go extinct within 10 generations. The dashed white line indicated parameters for which the variant has a 5% chance of surviving. The biological mean infectiousness (horizontal axis) has been scaled such that it equals the basic reproductive number (*R*_0_) in the homogeneous mixing scenario of panel A. For details on these calculations, see the Materials and Methods section.

In the unmitigated scenario (Fig. 1A), the procedure is relatively straightforward. A single infected individual is initially introduced, with a personal reproductive number *z* drawn from a negative binomial distribution *P*_NB_[*Z*; *R*_0_, *k*] with mean value *R*_0_ and dispersion parameter *k*. Thus, this individual gives rise to *z* new cases, and the algorithm is reiterated for each of these subsequent infections.

In the case of a lockdown scenario, in terms of restrictions of the number of social contacts (Fig. 1B), the algorithm is slightly more involved. In this case, a *degree c* (the number of contacts) is first drawn from a degree distribution (in this case a Poisson distribution, to mimic an Erdös-Renyi network). A biological reproductive number *ξ* (the *infectiousness*) is then drawn from a Gamma distribution with mean value *R*_0_ and dispersion parameter *k*. The actual personal reproductive number *z* is then drawn from the distribution

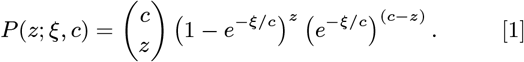

This reflects that the personal reproductive number *z* is, naturally enough, limited by the number of distinct social contacts *c*. This algorithms is then reiterated for each of the *z* new cases.

Similar results can be obtained analytically by considering the probability that an infection chain dies out in infinite time. Let that probability be *d* and let *p*_*i*_, *i* ∈ {0, 1, …} be the distribution of personal reproductive number (i.e. *p*_*i*_ is the probability that a single infected individual will infect *i* others). Then the extinction risk *d* is the sum:

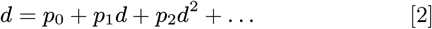

where the first term on the right hand side is the extinction risk due to the index case producing no new infections, the second term is the case where the index case gives rise to one branch of infections which then dies out (this being the reason for the single factor of *d* in the second term) and so on. Since each new branch exists independently of the other, the extinction events are independent and the probabilities may be combined by simple multiplication as in Eq. Eq. (2).

We find that the survival chance depends very strongly on overdispersion (Fig. 1), with more homogeneous variants (*k* ∼ 1) having a good chance of survival while highly overdispersed variants (*k* ≤ 0.1) are very unlikely to survive beyond 10 generations. This finding fits well with the general pattern of overdispersed spreading, namely that many individuals hardly become infectious while a few pass the disease onto many others. The uneven distribution of infectiousness makes heterogeneous diseases more fragile in the early stages of an epidemic, and thus more prone to stochastic extinction.

For the case of homogeneous mixing (Fig 1A) and the number of generations tending to infinity, Lloyd-Smith et al (6) performed a similar calculation using the generating function method described in Eq. 2. For a disease with *R*_0_ = 3 and a *k* value of 0.16 (similar to what they estimated for SARS-CoV-1), the survival chance was found to be 24%. Our model yields the same figure in the unmitigated connectivity→ ∞ limit.

To assess the effect of lockdown-like non-pharmaceutical interventions on the initial survival chances of a pathogen, we performed an analogous computation in a socially restricted setting (Fig. 1B). Compared with the unmitigated scenario of Fig. 1A, it can be seen that the mitigation has an effect on the survival chance, affecting highly overdispersed variants (small *k*) much more than their more homogeneous counterparts (with the same mean infectiousness). This result is parallel to the effect of lockdown-like interventions on the *competitive advantage* of a variant, which we explore in the next section.

In Ref. (20), the authors study stochastic extinction of a superspreading disease under a targeted intervention they call *cutting the tail*. They introduce a cutoff value *N*_cutoff_ for the personal reproductive number, and if a person has a personal reproductive number *z* ≥ *N*_cutoff_, a new *z* is drawn until one below the threshold is obtained. Since the disease is highly heterogeneous, this process is analogous to “removing” a potential superspreading event and replacing it with a much lower personal reproductive number (typically *z* = 0). This is exactly why the intervention is rightly called *targeted*. Their approach is thus based on viewing superspreading entirely as an event-based phenomenon, where one can directly remove superspreading events above some threshold size, and instead let the individuals take part in other less risky events. Our approach, on the other hand, assumes superspreading to be due to a combination of high individual biological infectiousness and opportunity, e.g. a large number of social contacts. These two viewpoints are complementary in obtaining a comprehensive description of superspreading phenomena, rather than mutually exclusive (17).

## Competitive advantage is determined by context

We now turn to the competition between two variants which have already managed to gain a foothold, and so have moved past the initial risk of stochastic extinction. This is a separate aspect of “fitness”, distinct from the initial survival ability described in the last section. Fig. 2 explores the competition between two strains which differ only in their level of overdispersion. The ancestral variant has a broad infectiousness distribution (*k* = 0.1) while the other – the *new variant* – is more narrowly distributed (*k* = 0.2). In the initial partial lockdown scenario, each person is only allowed contact with 10 others, At first, the fraction of infections due to the new variant is observed to grow rapidly. When it reaches a 20% share of active infections, around day 65, the lockdown is lifted (simulated by a shift to a homogeneous mixing contact structure). Naturally, this more permissive contact structure causes a surge in both variants (Fig. 2c). However, the fraction of infections owing to *each* variant suddenly stabilizes, indicating that the more homogeneous new variant has lost its competitive advantage in the unmitigated scenario.

**Fig. 2.**
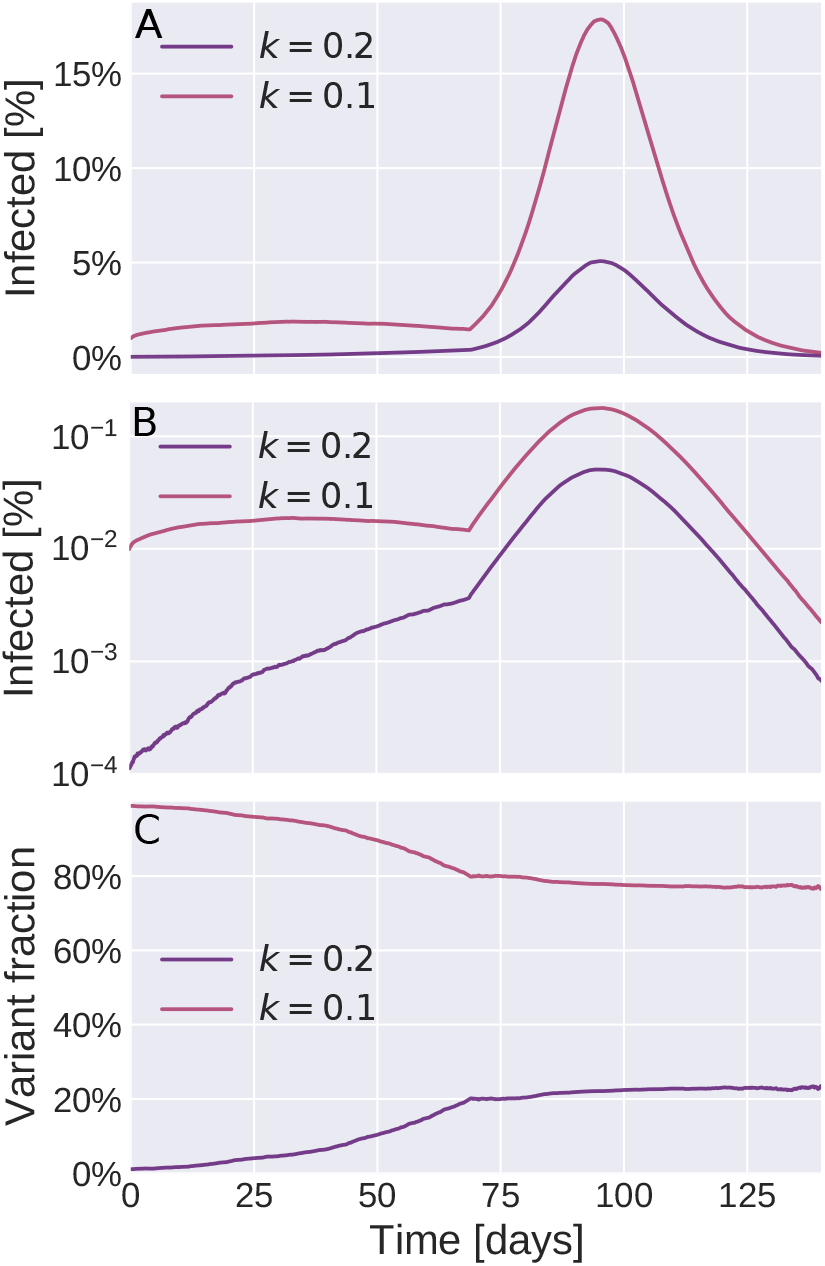
Simulations of the emergence of a new variant. An initially dominant (“ancestral”) strain with dispersion parameter *k* = 0.1 (red) has initially infected 1% of the population. The figure follows the emergence of a new variant (purple), which has the same biological mean infectiousness, but is more homogeneous (*k* = 0.2). Initially, 0.01% of the population is infected with the emerging variant. The two variants exhibit perfect cross-immunity. The initial scenario is a partially locked-down society (modeled as an Erdös-Renyi network with 10 contacts/person). When the new variant reaches 20% of all current infections (around day 65), the lockdown is completely lifted (modeled by a homogeneous mixing contact structure with the same *total* social time available per person). **A)** Incidence of each strain as a function of time since the new variant was introduced. Notice that the new variant spreads approximately exponentially until day 65 (see also panel B), whereas the ancestral strain stays at about 1% incidence. When restrictions are lifted, both surge. **B)** Same data as panel A, but plotted on a logarithmic scale. In this plot, exponential growth shows up as a straight line, and it is thus clear that the new variant spreads approximately exponentially during the lockdown phase. **C)** The relative proportions of the old and new variants. In the locked-down society, the new variant has a distinct fitness advantage, as revealed by its increasing share of infections. Once restrictions are lifted around *t* = 65 days, the fitness advantage is lost and the two variants spread equally well.

This sudden loss of competitive advantage demonstrates conceptually that the fitness of variants with different patterns of overdispersion depends on context, in the form of non-pharmaceutical interventions or the absence thereof. To quantify this dependence, we separately simulate the spread of several pathogen variants, each with its own specified mean infectiousness and dispersion parameter *k*, and measure the resulting basic reproductive numbers. In each case we let the pathogen spread in an Erdös-Renyi network with a mean connectivity of either 10 or 50, to simulate scenarios with either a restricted or fairly open society. The results are shown in Fig. 3, where the competitive (dis)advantage of each variant is plotted as a function of its a given biological mean infectiousness and dispersion. The infectiousness is given relative to the SARS-CoV-2 ancestral strain which is set to average infectiousness = 1 and has dispersion *k* = 0.1. This average infectiousness of 1 corresponds to a basic reproduction number of *R*_0_ = 3 in a well-mixed scenario, representative of COVID-19 (22). In the socially restricted case with only 10 contacts, the competitive advantage depends strongly on the dispersion parameter, as evidenced by the contour lines in Fig. 3A. The dashed white contour in the figure indicates variants which spread *as well* as the ancestral strain. Concretely, a variant with just half the biological infectiousness of the ancestral strain has no substantial competitive disadvantage, provided it is sufficiently homogeneous (*k* 1.0). In the more socially connected scenario (Fig. 3B), the competitiveness of a strain is observed to depend less strongly on dispersion, and is primarily determined by biological mean infectiousness. Viewed more broadly, these results imply that an observed increase in *R*_0_ for an emerging variant may be due to a *combination* of changes in transmission patterns (*k*) and biological mean infectiousness

**Fig. 3.**
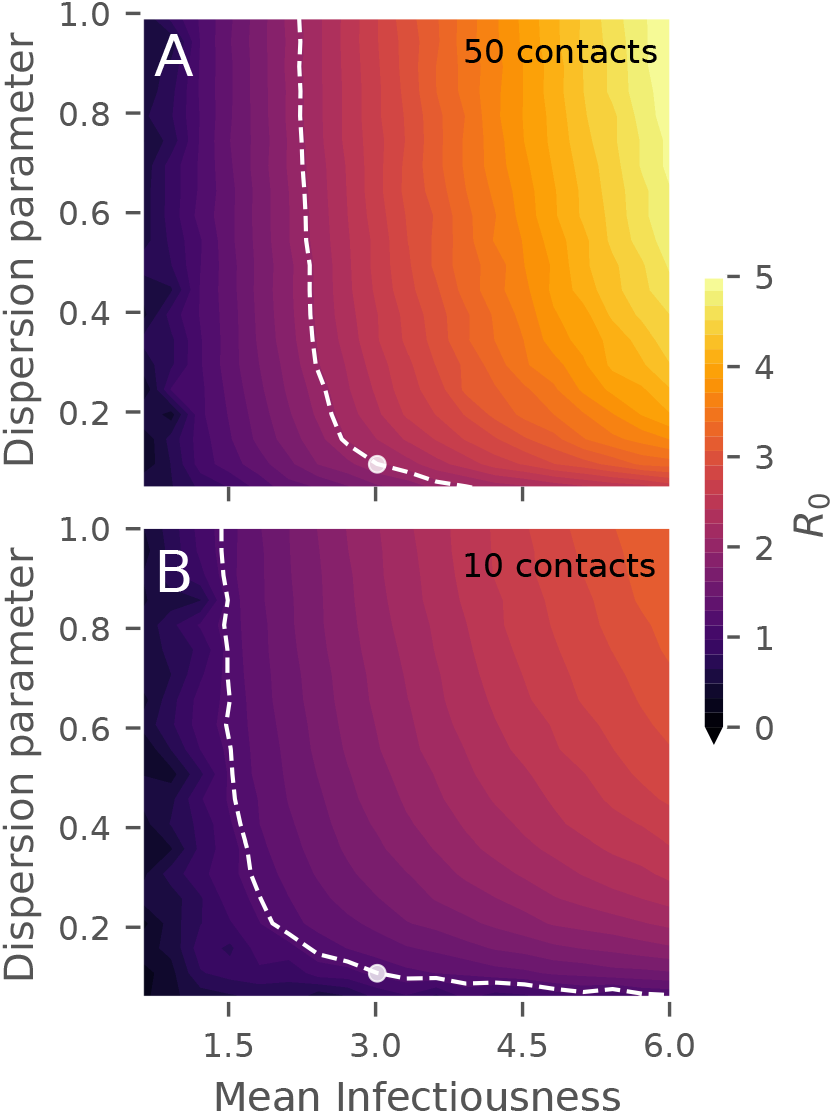
Relative fitness of variants. The color indicates the basic reproductive number that each variant exhibits under the given circumstances. The dashed white line indicates variants which have the same fitness as the ancestral strain, which is estimated to have *k* = 0.1. The biological mean infectiousness (horizontal axis) has been scaled such that it equals the basic reproductive number (*R*_0_) in a homogeneous mixing scenario. **A)** Spread of the disease in a connectivity 10 Erdös-Renyi network, corresponding to a partial lockdown. **B)** Spread of the disease in a connectivity 50 Erdös-Renyi network, corresponding to a mostly open society.

So far, our focus has been on mitigation strategies which rely on reductions in contact network. However, even when societies reopen by allowing contact with an increased number of individuals, non-pharmaceutical interventions which decrease transmission risk per encounter may be in force. These may include face masks and regular testing. In the Supporting Information, we show that interventions which decrease the transmission risk per encounter (i.e. per unit of contact time) in fact decrease the competitive advantage of more homogeneous variants. These types of interventions thus have essentially the opposite effect, relative to strategies which reduce social connectivity.

## Interventions exert selection pressure

As the observed differences in the viral load distributions of the Alpha (B.1.1.7.) variant and the ancestral strain suggest, overdispersion is not a fixed property, but rather one that may evolve over time. Furthermore, the SARS-CoV-2 pathogen has been estimated to mutate at a rate of approximately 2 substitutions per genome per month (23), translating to about one mutation per three transmissions. In Fig. 4, we explore the consequences of overdispersion as an evolving feature of the pathogen. In these simulations, the virus has a mutation probability of 1*/*3 at each transmission. When it mutates, the overdispersion factor is either increased (by a factor of 3*/*2) or decreased (by a factor of 2*/*3). Thus, we assume no drift on the microscopic scale, but one may arise macroscopically due to selection pressure from the environment. It should of course be noted that while the assumed mutation rate is realistic for SARS-CoV-2, many mutations will be neutral and only very few mutations will affect transmission dynamics. As such, the present model will likely overestimate the *magnitude* of the drift in overdispersion. It is however conceptually robust – decreasing the mutation rate merely slows down the drift, but the tendency remains.

**Fig. 4.**
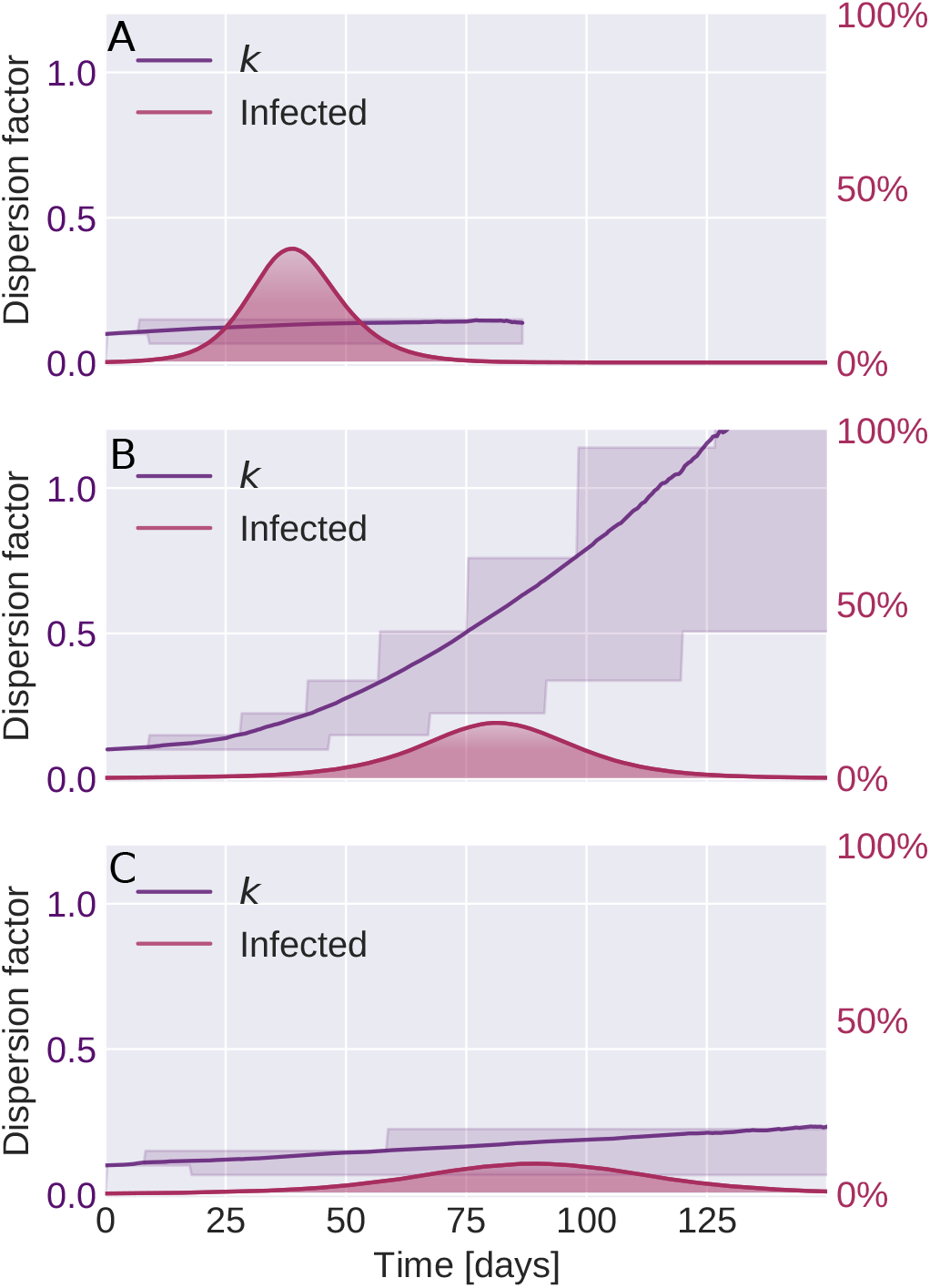
Evolution of overdispersion is driven by imposed restrictions. In these simulations, random mutations occur which alter the level of transmission overdispersion in a non-directed fashion. However, external evolutionary pressures are seen to drive the disease towards developing more homogeneous spreading patterns. The filled red curve shows the combined incidence of all strains. The purple curve shows the average dispersion factor *k* in the infected population (with higher *k* corresponding to a more homogeneous infectiousness). The shaded purple area shows the 25% and 75% percentiles of the distribution of dispersion factors in the infected population. **A)** The pathogen evolves in an open society with no restrictions imposed (homogeneous mixing contact structure). **B)** Partial lockdown, with an average social network connectivity restricted to 15 persons. **C)** No restrictions on social network, but infectiousness lowered by other means (e.g. face masks).

In our simulations, we find that there is always a tendency for overdispersion to decrease (i.e. for the *k* value to *increase*), leading to more homogeneous disease transmission. This makes sense, since we have already established that heterogeneous disease variants are more likely to undergo stochastic extinction (Fig. 1) and that they have a competitive disadvantage as soon as contact structures are anything but well-mixed (Fig. 3). In the absence of any interventions, the tendency to evolve towards homogeneity is quite weak (Fig. 4A), but when a partial lockdown is instituted, the picture changes dramatically and the *k* value increases exponentially. The conclusion is thus that lockdowns exert a selection pressure on the virus when it comes to overdispersion, towards developing a less superspreading-prone phenotype.

One may of course object that the scenarios of Fig. 4A (unrestricted spread) and 4B (partial lockdown) are not directly comparable, since the epidemic in 4A unfolds much more rapidly. For this reason, we have included the scenario shown in 4C, where the transmission rate per encounter has been lowered, but social structure is unrestricted. The transmission rate is lowered such that the *initial* daily growth rates in Fig. 4B and 4C are identical (11%*/*day averaged over the first 14 days). This slightly increases the growth of *k* over the course of the epidemic, but to a much lower level than in the lockdown scenario, demonstrating that it is indeed the restriction of social network that provides the selection pressure driving *k* upwards.

## Discussion

With this paper we have demonstrated that the relative success and survival of mutants of a superspreading disease depends on the type of mitigation strategies employed within a population. The choice of a certain mitigation strategy may well amount to selecting the next dominant variant. If, for example, a simple lockdown is enacted while still allowing people to meet within restricted social groups, the evolution of more homogeneously spreading disease variants may become favoured.

The spreading of an emerging virus in a human society is a complex phenomenon, where the actual reproductive number depends on sociocultural factors, mitigation policies and self-imposed changes in the behaviour of citizens as awareness grows in the population. The spread of a disease such as COVID-19 cannot simply be characterized by a single fitness quantity like the basic reproductive number *R*_0_, but will also depend on the heterogeneities of transmission patterns within the population. If schools are open, mutants which spread more easily among children may be selected for, whereas rapid self-isolation of infected individuals may tend to favor variants which temporally separate disease transmission from the development of symptoms. We have focused on modeling the evolutionary effects of biological superspreading in the context of mitigations such as lockdowns which have been implemented globally during the COVID-19 pandemic. We found that such lockdowns will favour the emergence of homogeneously spreading variants over time.

Our findings also have implications for the assessment of new variants. They highlight the importance of taking overdispersion into account when evaluating the transmissibility of an emerging variant. We have shown that the disease can spread more effectively not only by increasing its biological mean infectiousness, but also by changing its pattern of transmission to become more homogeneous. Practically, this means that transmission data obtained under even partial lockdown can lead to an overestimation of the transmissibility of an emerging variant. We thus call for an increased focus on measuring the overdispersion of variants, as this may be critical for estimating the reproductive number of new variants. These estimates in turn determine the required vaccination levels to reach herd immunity.

## Materials and Methods

We use an individual-based (or agent-based) network model of disease transmission as originally developed in Ref. (18). In this section, we present only a brief overview of the basic model, and refer to Ref. (18) for a more detailed description. We then go on to describe in detail the simulations and calculations which are particular to this manuscript.

The disease progression model consists of four overall states, **S**usceptible, **E**xposed, **I**nfected and **R**ecovered. The exposed state has an average duration of 2.4 days and is subdivided into two consecutive states with exponentially distributed waiting times (i.e. having constant probability rate for leaving the state) of 1.2 days each, thus constituting a gamma distributed state when viewed as a whole. The infectious state is divided into two states as well, of 1.2 and 5 days in duration, respectively.

Each individual in the model is associated with a fixed social network. Only a subset of edges are activated in each timestep, to simulate a contact event. In the simulations of this work, we always use either an Erdös-Renyi network with finite mean connectivity, or a homogeneous-mixing contact structure, which is also obtainable as the infinite connectivity limit of an Erdös-Renyi network.

When an edge connecting a susceptible and an infectious individual is active, there is a certain probability per unit of time for disease transmission to occur. This rate is determined by the individual infectiousness *r*_*i*_ of the infectious agent, which is drawn from a gamma distribution with dispersion parameter *k* before the individual has become infectious. As such, the infectiousness for any given individual is assumed constant throughout the infectious stage of the disease. The infectiousness distribution determines an upper bound on size Δ*t* of the the timesteps in the model, since the inequality *r*_*i*_ · Δ*t <* 1 must hold for all agents. A timestep of size Δ*t* = 30min was used throughout, since this was sufficient to ensure that the inequality was satisfied.

Below we go into more detail as to how the simulations involving multiple strains were performed.

### Stochastic extinction

The stochastic extinction (or, conversely, survival) plots of Figure 1 in the main text rely entirely on a branching process algorithm with sampling of probability distributions with an analytic description. In practice, we have performed the computation by numerical sampling.

In each generation of the epidemic, the computation is reiterated. Without loss of generality, we therefore here describe a single generation which initially has *I* infected individuals. Note that for the initial generation, *I* = 1 infected individuals.

- For *i* ∈ {1, …, *I*}:

- Draw individual infectiousness *ξ*_*i*_ from Gamma distribution *P*_*ξ*_(*ξ*; *k, µ*)
- Draw number of contacts *c* from a Poisson distribution with a given mean connectivity.
- Given number of contacts *c*, draw personal reproductive number *z*_*i*_ from the distribution Eq. (3)

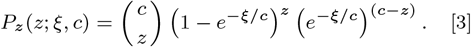

- Let the number of newly infected be *I =∑*_*I*_ *z*_*i*_ and repeat the algorithm with this new value of *I*.

If the number of infected *I* ever drops to zero, the outbreak is said to have undergone stochastic extinction in that generation. By performing multiple such branching process simulations for each value of the parameters *µ* (mean infectiousness) and *k* (dispersion factor) we build up a statistic of the survival chance of each specific variant. To generate Figure 1, this is repeated for two different values of the mean connectivity *c*.

### Two-strain competition simulations

In Fig. 2, two strains spread simultaneously in the population of *N* = 10^6^ individuals. Initially, 0.99% of the population are infected with the heterogeneous “old” variant (*k* = 0.1), while 0.01% are infected with the more homogeneous “new” variant (*k* = 0.2). Once a person with a given variant infects a susceptible individual, the characteristics of the variant are passed on to the newly infected individual, such that the infectiousness of this person is drawn from a Gamma distribution with dispersion parameter *k* set by the variant. In other words, these simulations assume that no further mutations affecting overdispersion occur, allowing us to track solely the competition of two differently-dispersed variants within a population.

### Evolutionary model

In Fig. 4, we allow the pathogen to stochastically mutate upon transmission, with the mutations affecting the degree of overdispersion. In the simulations, the pathogen mutates on average once for each new host it is transmitted to (i.e. with mutation probability *p* = 1*/*3) and the mutations are assumed to always affect overdispersion, by either increasing the *k* value by a factor of 3*/*2 (i.e. *k* → 3*k/*2) or decreasing it by a factor of 2*/*3 (i.e. *k* → 2*k/*3). On a microscopic level, the dispersion level thus performs an unbiased (multiplicative) random walk. The value of this step-size parameter is arbitrarily chosen, and as such the simulations can only be regarded as qualitative and conceptual. However, although no intrinsic bias is built into the mutation mechanism, external selection pressures may drive the level of overdispersion in the population up or down, as is explored in Fig. 4.

In Fig. 4C, the average infectiousness of the strain is lowered so as to produce an initial growth rate that is identical to that of 4A, namely 11% per day in the first 14 days of the epidemic.

## Data Availability

Data (simulation code) is available in a public, DOI-enabled GitHub repository.

https://github.com/BjarkeFN/OverdispEvolve

## ACKNOWLEDGMENTS

We thank Robert J. Taylor, and Julius B. Kirkegaard for enlightening discussions. Our research has received funding from the European Research Council (ERC) under the European Union’s Horizon 2020 research and innovation programme, grant agreement No. 740704, as well as from the Carlsberg Foundation under its Semper Ardens programme (grant # CF20-0046).

