## Supporting Information for "Lockdowns exert selection pressure on overdispersion of SARS-CoV-2 variants"

### Supporting Information: Lockdowns exert selection pressure through overdispersion of SARS-CoV-2 variants

(Dated: June 30, 2021)

#### INTERVENTIONS TARGETING TRANSMISSION RISK

In the main text we have primarily considered the evolutionary pressures exerted by mitigation strategies which rely on reductions in social connectivity, such as lockdowns. However, other mitigation strategies may rely not on limiting the number of distinct individuals that a person interacts with, but rather on decreasing the transmission rate when contacts *do* occur. Examples of such strategies include the use of face masks, which work primarily by decreasing the number of emitted virions, and physical distancing which exploits the decrease in viral concentration as distance to the source is increased. In Fig. 1, we consider two scenarios in which a more homogeneous variant with dispersion factor $k = 0.2$ emerges in the background of the $k = 0.1$ ancestral strain. Note that the curves in Fig. 1 show the relative *share* of infections owing to the emerging variant and not the absolute incidence. In both of the cases studied, social connectivity is initially quite restrictive, with each person allowed only 10 contacts. At time $t = 25$days, restrictions are relaxed, simulated by increasing average connectivity to 50. This is where the two scenarios diverge: in one of them, the infection risk *per encounter* is halved, while in the other it stays the same. It is of course expected that halving the infection risk will cause a lower overall epidemic, but what is not obvious is how it will affect the competition between the two variants. As expected from Fig. 3 of the main text, the competitive advantage of the new variant is reduced by going from 10 to 50 social connections, as is reflected in both of the curves in Fig. 1 bending off at $t = 25$days. However, more notable is the fact that the competitive advantage of the new variant is substantially reduced when restrictions are put in place which decrease the infection rate. The conclusion is thus that heterogeneous variants are particularly vulnerable to lockdown-type interventions, where social network size is reduced, while more homogeneous variants are more susceptible to interventions which reduce infectiousness during each encounter.

In Fig. 2 we show the absolute incidence of the two variants in another simulation experiment where the two types of interventions are in force simultaneously. Initially, only the (partial) lockdown is in force, with social connectivity restricted to 10 persons. At $t = 35$ days, another non-pharmaceutical intervention which reduces the transmission risk per encounter by 20% is put in force. This is seen to reduce the more homogeneous emerging variant to marginal spread while the ancestral strain starts to decline.

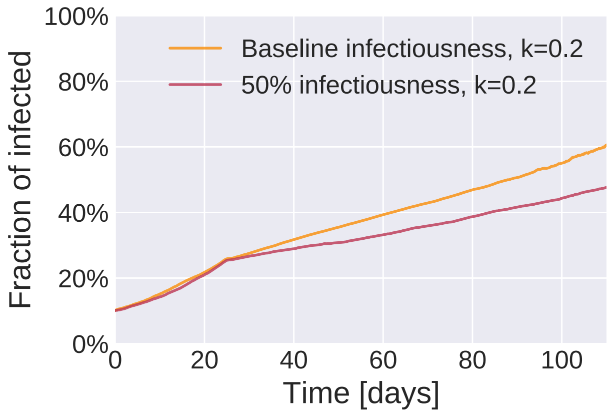


FIG. 1. **Re-opening society while putting other restrictions in place.** The plot shows the relative abundance of a new, more homogeneous variant ($k = 0.2$) as a function of time. At time $t = 0$, the new variant makes up 10% of infections, while the rest owe to the more heterogeneous ”old” variant ($k = 0.1$). Until time $t = 25$ a partial lockdown is in place, modeled by restricting personal contact networks to only 10 persons on average. At $t = 25$days, society is partially opened, simulated by allowing contact with 50 different persons. The red line represents a scenario where the opening of society is accompanied by other restrictions, reducing the infection risk *per encounter* by half. As such, the diversity of contacts encountered is increased, but the infection risk per encounter is decreased. In the scenario shown by the yellow line, infection risk per encounter is unaltered. Clearly, these interventions negatively affect the competitive advantage of the more homogeneous variant.

*
†
‡
§

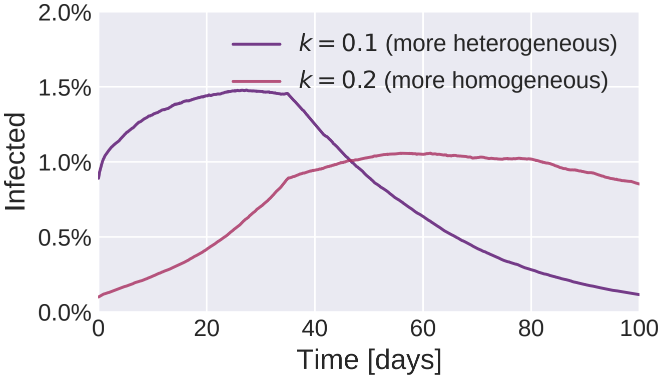


FIG. 2. Two variants with dispersion parameters $k = 0.1$ and $k = 0.2$ spread in a socially restricted society with a personal contact network size of 10. From time $t = 35$days, non-pharmaceutical interventions are introduced which reduce the rate of transmission by 20%. This is enough to reduce the more homogeneous ($k = 0.2$) variant to marginal growth, while the heterogeneous variant dies out.
